# Patient-specific compliant simulation framework informed by 4DMRI-extracted Pulse Wave Velocity: Application post-TEVAR

**DOI:** 10.1101/2024.03.17.24304341

**Authors:** Louis Girardin, Niklas Lind, Hendrik von Tengg-Kobligk, Stavroula Balabani, Vanessa Díaz-Zuccarini

## Abstract

We introduce a new computational framework that makes use of the Pulse Wave Velocity (PWV) extracted exclusively from 4D flow MRI (4DMRI) to inform patient-specific compliant computational fluid dynamics (CFD) simulations of a Type-B aortic dissection (TBAD), post-thoracic endovascular aortic repair (TEVAR). From 4DMRI and brachial pressure, a 3D inlet velocity profile (IVP), dynamic outlet boundary, and reconstructed thoracic aortic geometry are obtained. A moving boundary method (MBM) is applied to simulate aortic wall displacement. The aortic wall stiffness was estimated through two methods: one relying on area-based distensibility and the other utilising regional pulse wave velocity (RPWV) distensibility, further fine-tuned to align with *in vivo* values. Predicted pressures and outlet flow rates were within 2.3% of target values. RPWV-based simulations were more accurate in replicating *in vivo* hemodynamic compared to the area-based ones. RPWVs were closely predicted in most regions, with the exception being the endograft, and systolic flow reversal ratios (SFRR) were accurately captured, while a difference of above 60% on in-plane rotational flow (IRF) between the simulations. Significant disparities between the wall shear stress (WSS)-based indices were observed between the two approaches, especially the endothelial cell activation potential (ECAP). At the isthmus, the RPWV-driven simulation indicated a mean ECAP>1.4*Pa*^−1^ (critical threshold), indicating areas potentially prone to thrombosis. In contrast, the area-based simulation did not depict this. RPWV-driven simulation results agree well with 4DMRI measurements, emphasising that RPWV simulations are accurate in simulating haemodynamics, consequently facilitating a comprehensive assessment of surgery decision-making and potential complications, such as thrombosis and aortic growth.

## 1. Introduction

Type-B Aortic Dissection (TBAD) affects 3 in 100,000 people annually (Trahanas et al., 2022).Surgical intervention is necessary when complications like aneurysmal dilatation arise. TBAD open surgery carries high mortality rates, about 20% (Yuan et al., 2018). In recent decades, thoracic endovascular aortic repair (TEVAR) has emerged as a less invasive surgical approach with favourable post-operative survival rates (Jiang et al., 2023). TEVAR is an endovascular surgery which aims to restore normal aortic function by covering the initial entry tear and promoting thrombosis of the false lumen (Uchida and Sadahiro, 2018). While various studies have highlighted TEVAR advantages (Nienaber et al., 2013; Singh et al., 2021; Tadros et al., 2019), TEVAR remains associated with outcomes such as endoleaks and high mortality rates (Bavaria et al., 2022). TEVAR for TBAD requires follow-up screenings to prevent and predict complications (Eidt and Vasquez, 2023; Williams et al., 2022).

Elevated pulse wave velocity (PWV) is associated with aortic stiffening and TEVAR, as the endograft rigidifies the aorta (De Beaufort et al., 2017; Hori et al., 2020). Increasing PWV has been linked with cardiovascular outcomes such as stroke and left ventricular hypertrophy (Valencia-Hernández et al., 2022). PWV can be determined through 2D flow MRI or cine-MRI by assessing the pulse wave travel time along the aortic centerline or the distensibility by measuring the cross-sectional variations of the vessel throughout the cardiac cycle (Wentland et al., 2014). The regional PWV (RPWV) can be measured on 4DMRI, providing the local stiffness of the vessel (Nguyen et al., 2023; Wentland et al., 2014). RPWV is significant in the context of TEVAR due to the proximal aortic stiffening after the endograft placement (Bissacco et al., 2022).

TEVAR can induce aortic wall remodelling and abnormal flow, affecting wall shear stress (WSS) (Midulla et al., 2021), possibly leading to endothelial dysfunction with outcomes such as thrombosis (Morbiducci et al., 2009; Nauta et al., 2017). 4DMRI allows the assessment of various functional parameters, such as blood flow dynamics, which is not conventionally measured in the clinical assessment of TEVAR (Cosset et al., 2022; Takahashi et al., 2022). However, its low near-wall spatial resolution limits the quantification of pressure and WSS in abnormal regions (Lamata et al., 2014; Markl et al., 2011). Computational fluid dynamics (CFD) simulations can be fused with 4DMRI to calculate these haemodynamic variables accurately (Armour et al., 2022; Black et al., 2023).

CFD simulations are beneficial in the context of TEVAR to assess the impact of compliance mismatch introduced by endografts. Fluid-structure interaction (FSI) has traditionally been used for patient-specific simulations of TEVAR procedures, for example, to study their impact on cardiac remodelling and left ventricular afterload (Van Bakel et al., 2019) or the flow reversal due to the rigidity and length of the endograft (Aghilinejad et al., 2022) and their impact on left ventricular afterload. However, FSI is computationally costly and often based on literature values of elastic or anisotropic tissue data, as *in vivo* data are barely accessible (Wang et al., 2023) or impossible to obtain. To circumvent these challenges, different alternatives have been developed. Recently, a mesh morphing technique based on dynamic CT imaging was proposed to allow the reconstruction of the transient aortic geometry(Capellini et al., 2021, 2018). The technique was used to study large-scale flow coherence in ascending aorta(Calò et al., 2023). A moving boundary method (MBM) was developed, in-house, as a less-costly alternative based on a non-imposed radial nodal displacement calculation and has been applied in several studies (Bonfanti et al., 2018, 2017; Girardin et al., 2024; Stokes et al., 2021).

In this study, a 4DMRI-informed computational method based on our MBM is developed, using 4D flow MR and non-invasive pressure measurements to inform patient-specific, compliant CFD simulations of TBAD post-TEVAR. More specifically, RPWVs, a routinely used clinical marker, are extracted from 4DMRI using a cross-correlation method to tune iteratively the aortic stiffness used in the simulations. Additional simulations with stiffness derived from the area-based distensibility are conducted. Hemodynamic indices are estimated and compared.

## 2. Methods

### 2.1. Data Acquisition

A patient with chronic TBAD previously treated with TEVAR was presented for follow-up at Inselspital Bern. Their aorta was imaged following an ethically approved protocol (Local Institutional Review Board ID 2019-00556). MRI sequences of the thoracic aorta down to the abdominal aorta were acquired using a MAGNETOM Sola fit scanner (Siemens Healthineers). 4DMRI was acquired with a resolution of 2.5 mm*2.5 mm*2.5 mm. A T2/T1 weighted TRUFI MRI sequence was acquired with a 1 mm*1 mm* 1 mm resolution. Cine-MRI and 2D flow MRI (2DMRI) were acquired with a resolution of 1.875 mm*1.875 mm. Brachial pressure was acquired before the examinations.

### 2.2. Segmentations and Meshing

The aorta was segmented from the T2/T1 TRUFI using ScanIP (Synopsis Simpleware, USA). The combined geometry was smoothed using MeshMixer (Autodesk, USA). Inlet and outlets were trimmed perpendicularly to their cross-sectional area using Fluent Mesh (Ansys Fluent, USA) (Fig 1a). The geometry was separated into discrete regions along the centreline based on anatomical and stiffness characteristics observed with the wall displacement features on the 4DMRI (Fig 1).

**Fig 1.**
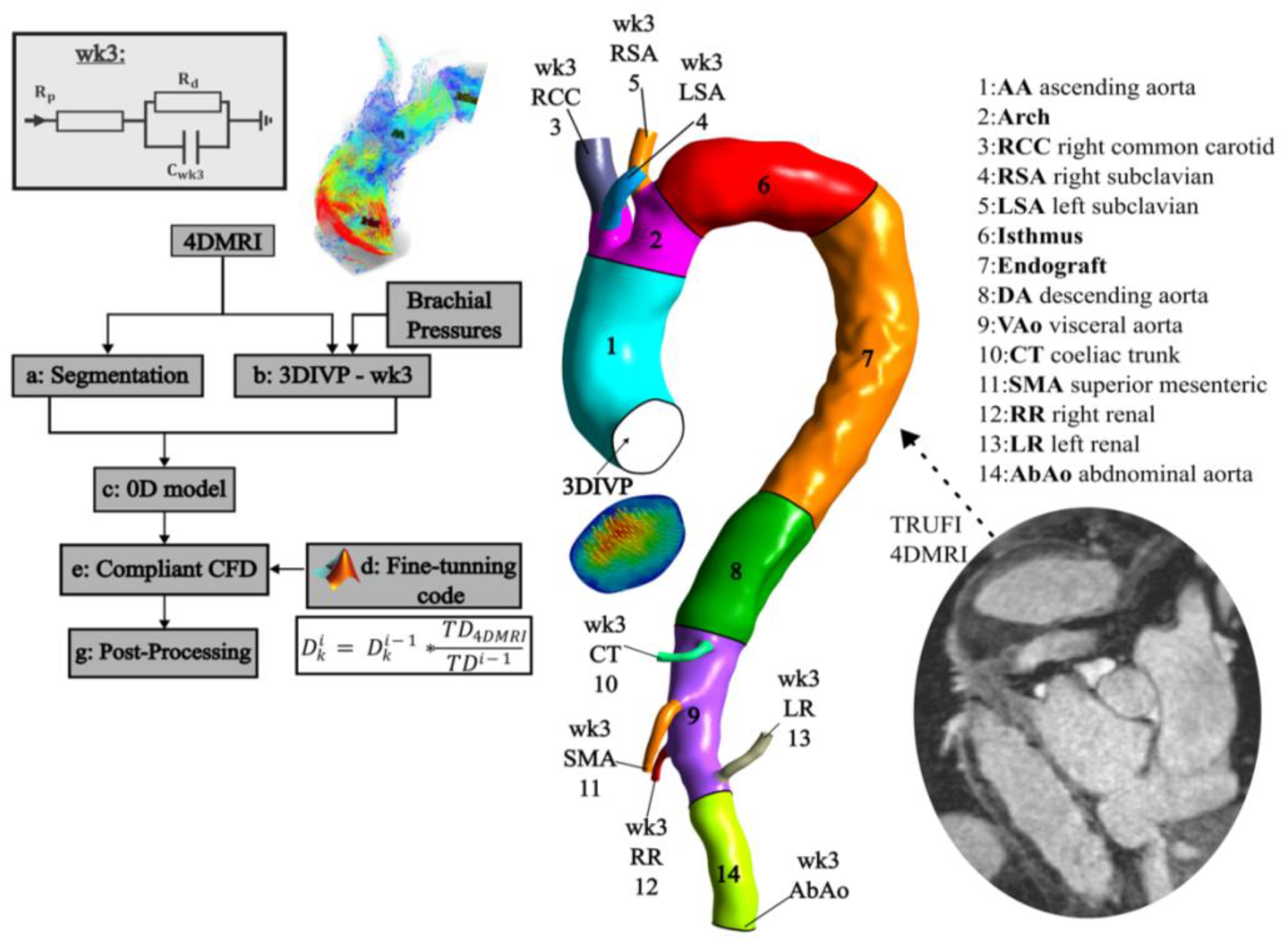
Process diagram showing the calibration steps (a-g) for the patient-specific compliant simulation, informed via RPWVs.

A tetrahedral computational mesh was created using the segmented geometry with Fluent Mesh 19.0 (Ansys Inc., USA). Details on the mesh element sizing, parameters used for the prism layering, and the mesh sensitivity analysis used to obtain the final mesh are available in Appendix 1.

### 2.3. Boundary Conditions

Following our previous work (Stokes et al., 2023b), 4DMRI was used to extract a 3D inlet velocity profile (3DIVP) and outlet mean flow rates (Fig. 1a) using GTFlow (GyroTools LLC, Switzerland) (Table 1). The 3DIVP was spline-interpolated to apply a 1ms CFD timestep using MATLAB (MathWorks Inc., USA) (Fig. 1b).

**Table 1.** Target mean flow rates at outflow boundaries and inlet systolic and diastolic pressures against CFD values.

|  | Flow rate [mL/s] |  |  |  |  |  |  |  | Pressure [mmHg] |  |
| --- | --- | --- | --- | --- | --- | --- | --- | --- | --- | --- |
|  | RCC | RSA | LCC | CT | SMA | LR | RR | AbAo | Systole | Diastole |
| **Target** | 18.88 | 7.41 | 8.12 | 3.13 | 8.25 | 1.87 | 1.50 | 14.70 | 101.8 | 70 |
| **S1** | 18.98 | 7.38 | 8.09 | 3.17 | 8.21 | 1.89 | 1.50 | 14.63 | 100.36 | 68.4 |
| **S2** | 18.75 | 7.35 | 8.15 | 3.12 | 8.33 | 1.84 | 1.53 | 14.75 | 102.5 | 70.5 |

A 0D lumped parameter model of the vasculature (20-sim, Controllab Products, Netherlands) was used to calibrate three-element Windkessel (WK3) parameters (Fig 1c), used at the outlets as pressure boundary conditions in the CFD as described in previous works (Stokes et al., 2023b; Westerhof et al., 2009). Parameter values are displayed in Appendix 2.

### 2.4. Moving Boundary Method & Stiffness Tuning

The MBM introduced by (Bonfanti et al., 2018)was used to simulate the compliant behaviour of the aorta. According to this method, the local wall displacement follows the surface node normal, proportional to the difference between nodal pressure and exterior pressure and the local wall stiffness. The stiffness *K*_*i*_ (N/m^3^) is equal to:

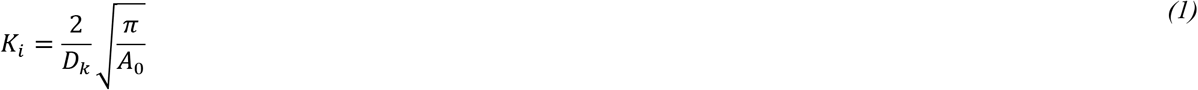

where *A*_0_ (m^2^) is the local cross-sectional area at diastole, and *D*_*k*_ (1/Pa) is the local wall distensibility. Local stiffness values (*K*_*i*_) were initially calculated using a distensibility *D*_*k*_ defined by the ratio between the cross-sectional relative change, as measured by the 4DMR images, and the regional pulse pressure, such as:

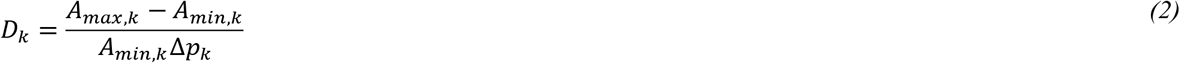

where *A*_*max,k*_ and *A*_*min,k*_ (m^2^) are the maximum and minimum cross-sectional area of the aortic vessel in a region *k* and Δ*p*_*k*_ is the pulse pressure in the region *k*, estimated through transient CFD simulations assuming a rigid wall. When no displacements could be extracted from 4DMRI due to its spatial resolution, the distensibility was estimated from the RPWV using the empirical relationship from (Reymond et al., 2009):

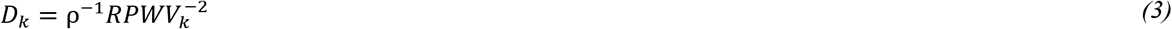

Where *ρ* is the density of blood (*kg/m*^3^). RPWVs were estimated from the 4DMRI in each region using a cross-correlation method (Fielden et al., 2008; Markl et al., 2010), as illustrated in Fig. 2.

**Fig 2.**
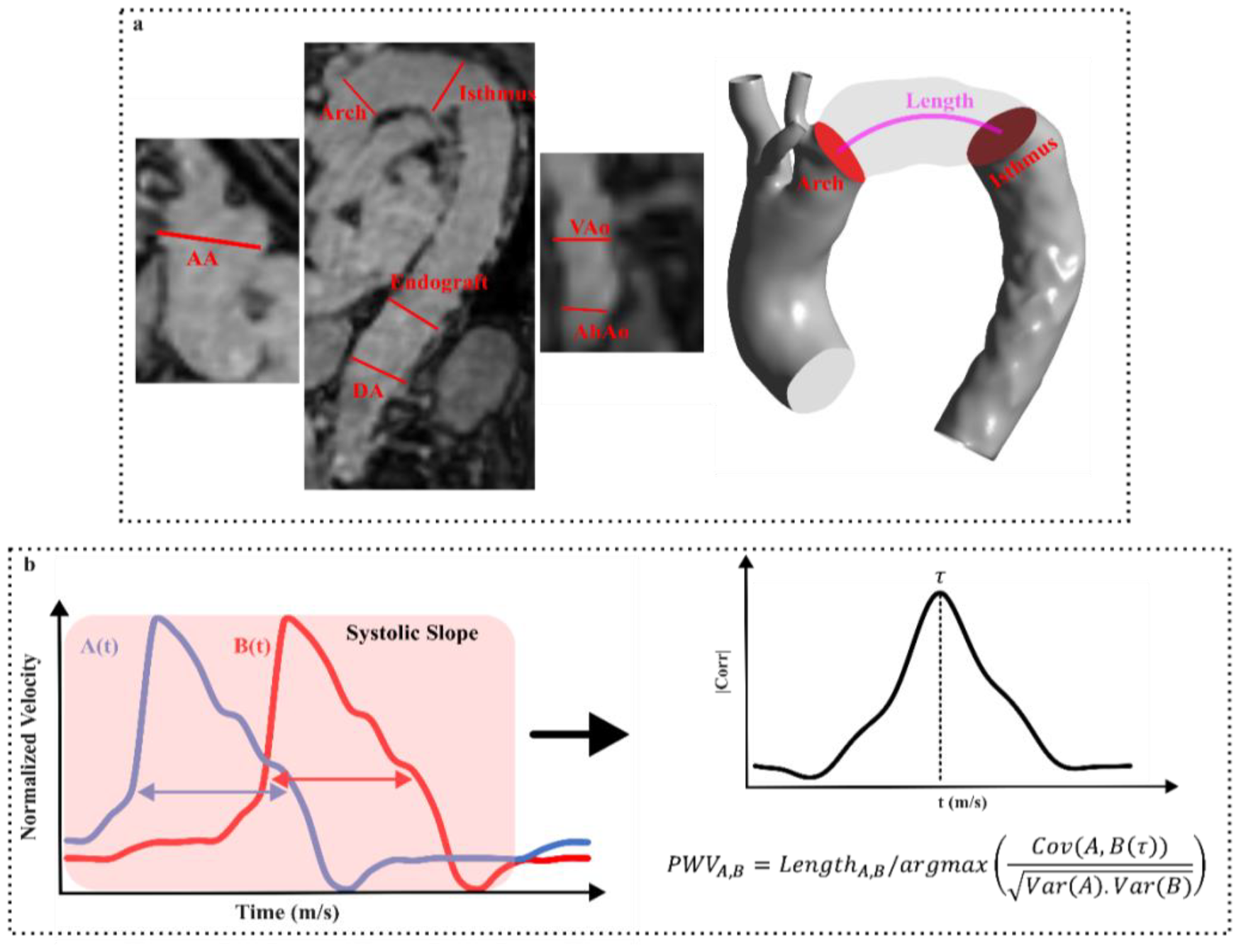
a.- Red lines depict the 4DRMI planes on which the flow rates were measured. b.- schematic of the cross-correlation method used to calculate the PWV between planes A and B.

In the endograft region, this method yielded a distensibility of 1.6 * 10^−3^*mmHg*^−1^, which is an order of magnitude higher than reported values in the literature (Johnston et al., 2010; Rovas et al., 2023; Tremblay et al., 2009). Considering the possibility of measurement noise proximal to the endograft, we opted to use a literature-based distensibility value of 1.6 * 10^−4^*mmHg*^−1^ in the calculation of stiffness in this region.

A varying, patient-specific stiffness field was thus calculated using RPWV-based distensibility. An automatic iterative method was developed to tune the stiffness field to match the 4DMRI RPWVs and exemplified in Fig. 1. A MATLAB code calls CFX (Ansys Inc., USA) and updates the simulation script with a new stiffness in each run. Only the systolic phase was run to avoid potential wave reflection during diastole and to reduce the computational time of the iterative process. At the end of each run, flow rates were extracted from the planes enclosing the regions of interest and the RPWVs were calculated (Fig 2). The distensibility of each region *k, D*_*k*_ (1/Pa), was then updated following the equation below:

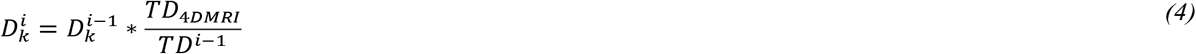

The process was run until the time delay error was lower than 10%; seven iterations were required, each taking 3 hours in an 8 Intel Core i9-11900K. The final stiffness map was inputted to the full simulation (Fig 3). S1 denotes simulations obtained with the stiffness map extracted from the area changes (equations 1 and 2), and S2 is the one based on RPWV estimates, using equations 1 and 4.

**Fig 3.**
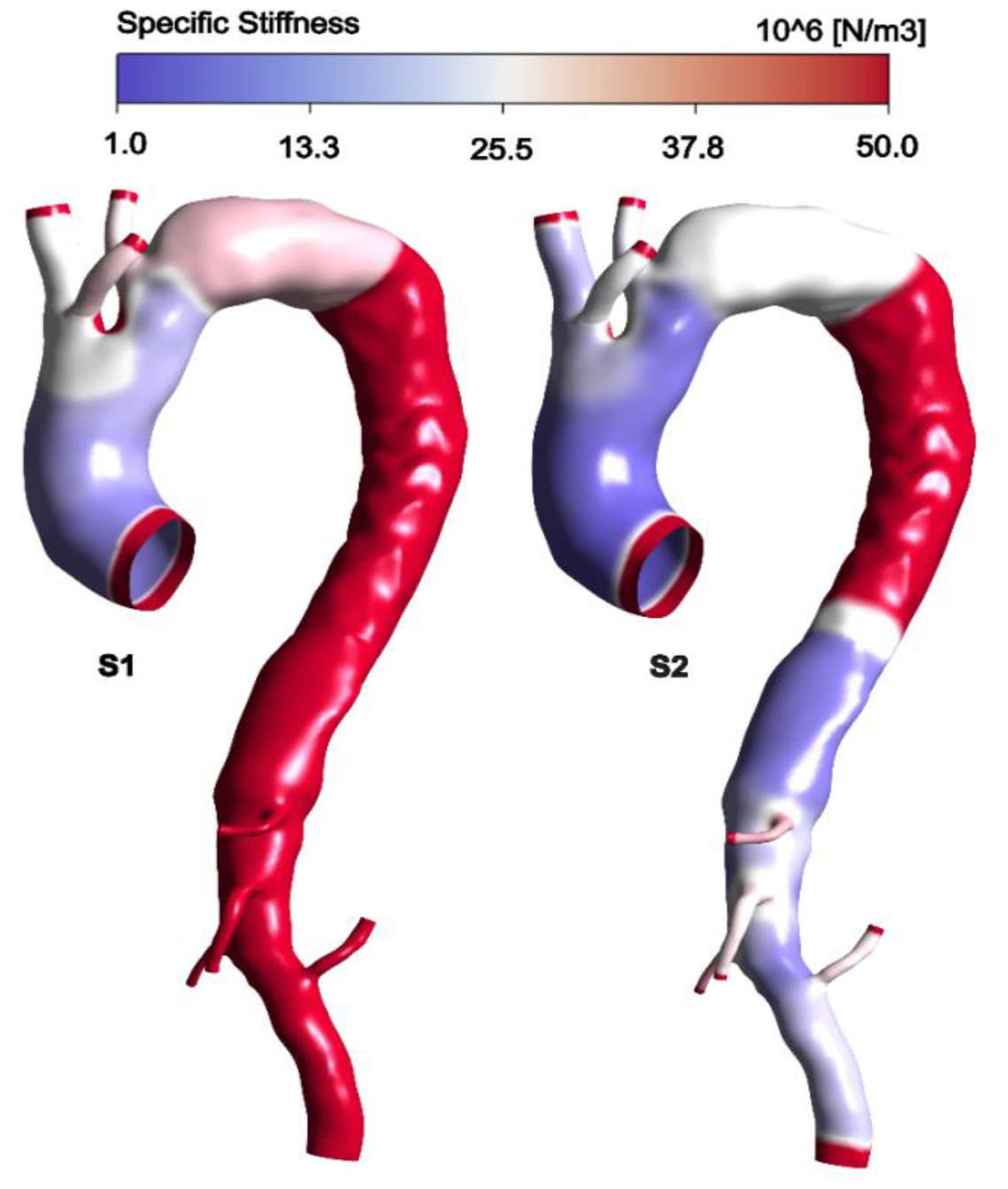
Stiffness maps produced from area- and RPWV-based distensibility used to compare S1 and S2 with a logarithmic scale, respectively.

### 2.5. Simulation Model

The finite-volume solver ANSYS CFX 19.0 was employed to solve the transient three-dimensional Navier-Stokes equations (Fig. 1d,e). These equations considered the Carreau-Yasuda viscosity model and used empirical constants derived from the work of (Tomaiuolo et al., 2016). Blood was modelled as incompressible and non-Newtonian, with a 1056 kg/m^3^ density. The peak *Re*_*p*_ and critical *Re*_*c*_ Reynolds numbers computed as in (Peacock et al., 1998), were equal to 6855 and 6000, respectively. Therefore, the k-ω shear stress transport model was employed to model the turbulence with a low turbulence intensity of 1% introduced to the flow (Kousera et al., 2013). The Navier-Stokes and continuity equations were solved with a second-order backward Euler scheme, using a 1ms time step. The convergence criterion was set to a root-mean-square residual value of 10^−5^ for all equations within each time step. A state of periodicity characterised by less than 1% variation in systolic and diastolic pressures between cycles (Fig. 1d,g) was reached after four cycles for both simulations, and the results of the last cycle were post-processed to extract the hemodynamic indices reported below.

## 3. Results

### 3.1. Validation

Mean flow rates and diastolic and systolic pressures are shown against target values in Table 1. S1 and S2 pressures and mean flow rates at flow boundaries were predicted within 2.3% of error.

The simulated RPWVs were compared against the 4DMRI ones in the regions of interest (Fig 4). Results show that S1 systematically overestimates PWV due to the high stiffness values across most of the flow domain, starting from the isthmus to the AbAo (Fig 3). In S2, a maximum error of 8% was found in the arch and DA regions except the endograft; this corresponds to a 1 ms difference in travel time.

**Fig 4.**
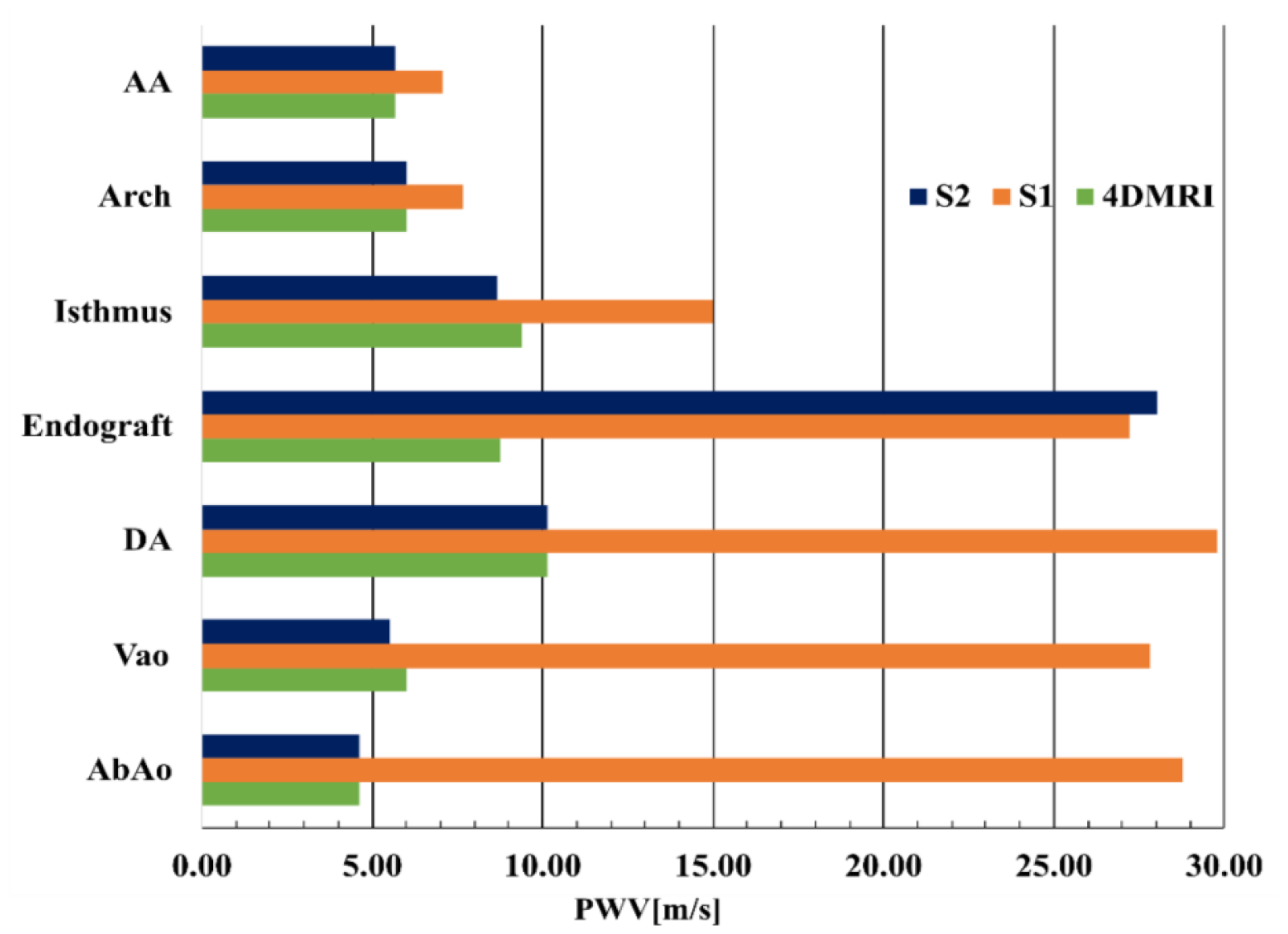
Comparison of predicted RPWVs, S1 (orange) and S2 (green), against 4DMRI extracted (blue).

### 3.2. Flow characteristics

The systolic flow reversal ratio (SFRR), the ratio between backward and forward flow during systole, is a clinical marker linked with local dilation when increasing (Gil-Sala et al., 2021). SFRR was evaluated across the planes enclosing the regions of interest shown in Fig 1a. 4DMRI estimated SFRR values exceeded 30% in the AA region (Fig 5), which can be attributed to the circulating flow pattern at the inlet. Moreover, the stenosis at the arch causes upstream flow acceleration and recirculation and a build-up of pressure before the stenosis, leading to about half of the flow going backwards during systole. The compliance mismatch between the proximal aortic aorta and the endograft can lead to a pronounced SFRR (Sultan et al., 2021). In our study, an SFRR >30% at the interface between the isthmus and the DA with the endograft is consistent with these reported findings. Downstream of the endograft, where the compliance mismatch is less pronounced and the geometry more tubular and straight, the SFRR decreases to 18% and 14% at the VAo and AbAo, respectively. The maximum difference between the 4DMRI and the predicted SFRR in S1 occurred at the DA.

**Fig 5.**
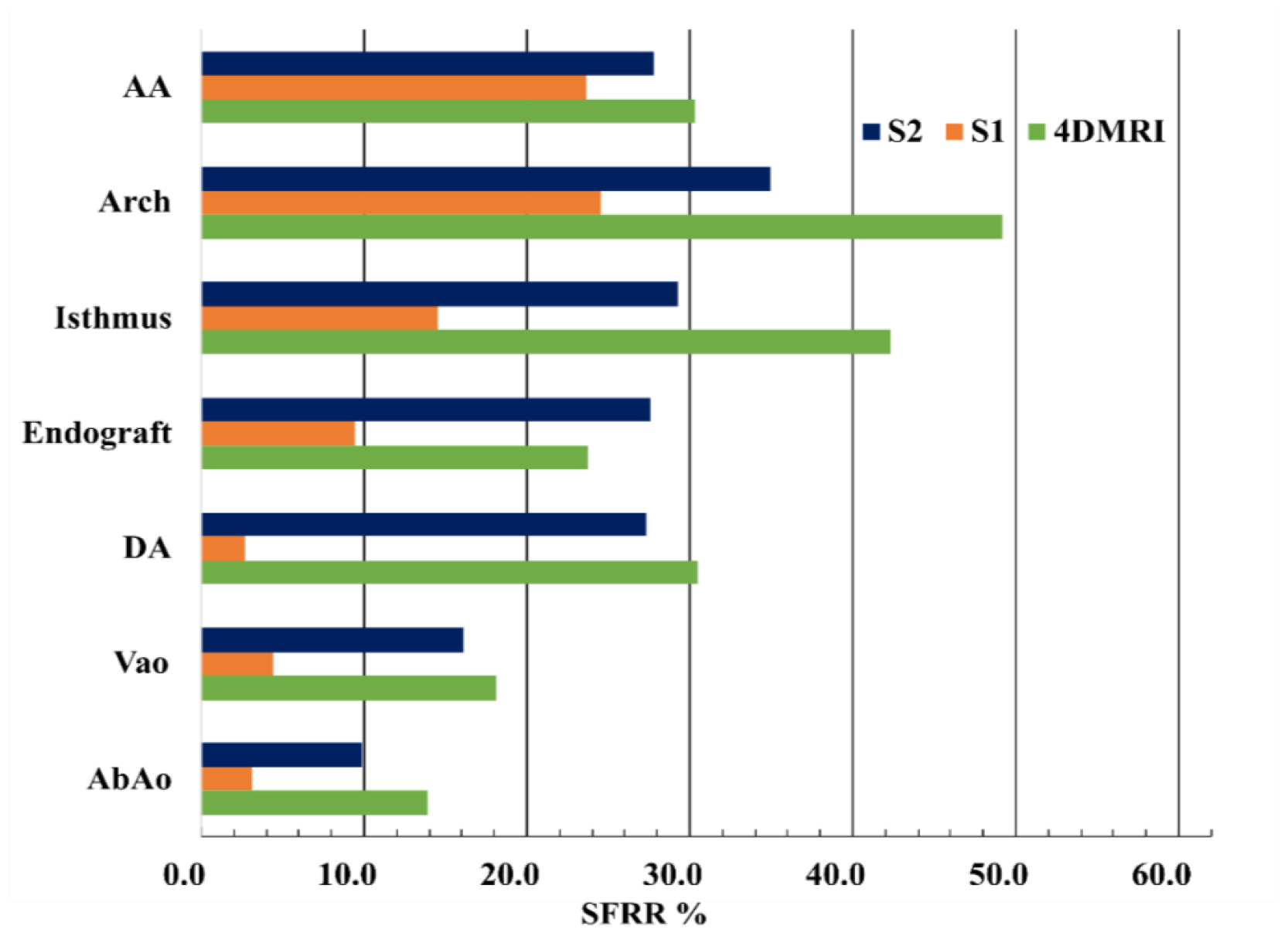
Comparison between 4DMRI (blue) measured and predicted -, S1 (orange) and S2 (green)-SFRR values along the aortic domain.

Previous research showed the reduction of the in-plane rotational flow (IRF) in rigidified and dilated aortae, which are correlated outcomes of TBAD and TEVAR (Dux-Santoy et al., 2019; Gil-Sala et al., 2021). IRF is not presented for the 4DMRI due to the spatial resolution limitations, resulting in inaccurate calculations of the finite differences used to evaluate the vorticity. Similarly to the flow reversal trends, S1 appear to overpredict IRF compared to S2, particularly in regions with significant stiffness differences between the two simulations, such as at the isthmus, DA, VAo and AbAo, with differences exceeding 60% (Fig. 6).

**Fig 6.**
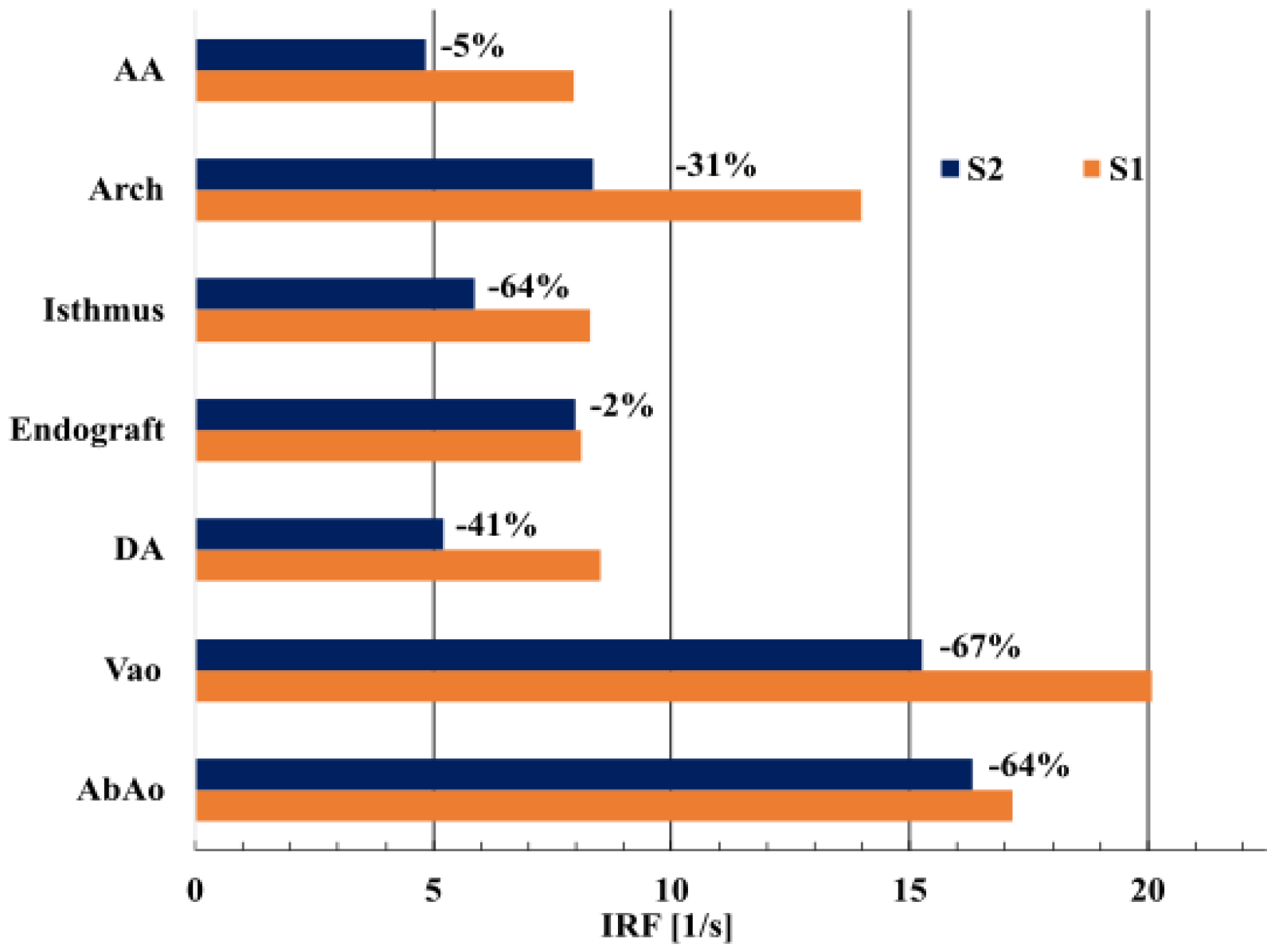
IRF for S1 (blue) and S2 (orange) with the difference reported next to the bar.

Velocity magnitude contours at different flow phases and on selected planes are shown in Fig. 7. S2 demonstrates a better qualitative agreement with the 4DMRI, capturing velocity magnitude distribution more accurately, such as at T1 and T3 on ii. The peak velocity displacement is also better reproduced in S2, such as at T3 on i and ii. A limitation observed in the 4DMRI data is the resulting coarser images at T4 during late diastole, which makes comparison with simulations difficult, and which a priori show limited agreement likely due to poor signal to noise ratio.

**Fig 7.**
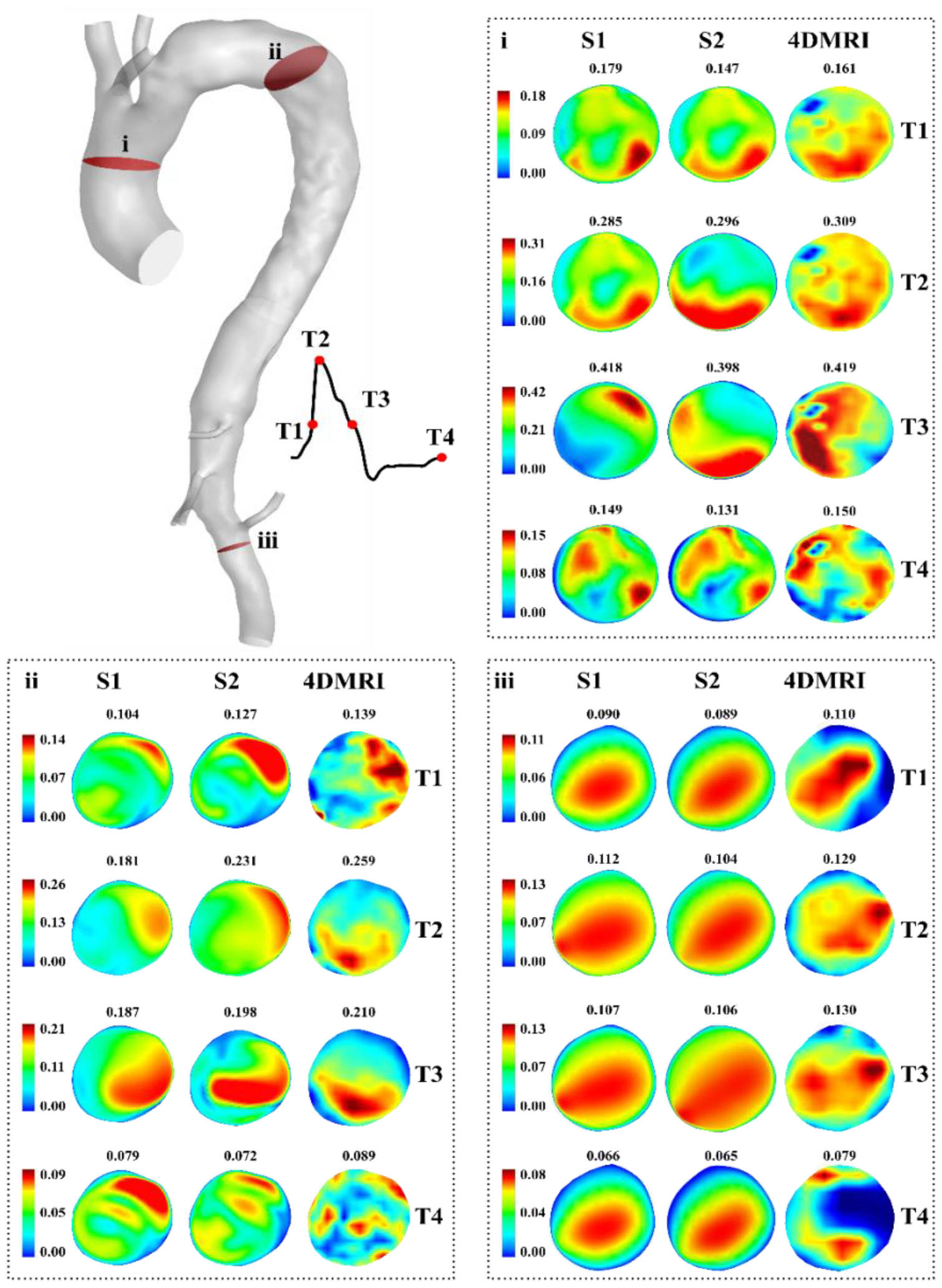
Contours of velocity magnitude at different flow phases in each case at the AA (i), endograft (ii) and Vao (iii).

### 3.3. Wall shear stress indices

Contours of time-average wall shear stress (TAWSS), oscillatory shear index (OSI) and endothelial cell activation potential (ECAP) (Di Achille et al., 2014; D. Gallo et al., 2012) predicted from the two simulation approaches and the point-wise difference between them are displayed in Fig 8. Table 2 displays the minimum, maximum and mean values on each region for S2 and the average relative error between S2 and S1 per region.

**Table 2.** Minimum, maximum and mean values per region for S2. Also shown are mean relative errors value such as 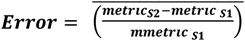.

|  |  | **TAWSS**<br>[Pa] | **OSI**<br>- | **ECAP**<br>[Pa] |
| --- | --- | --- | --- | --- |
| **1:AA** | Min | 0.26 | 0.01 | 0.00 |
|  | Max | 1.83 | 0.49 | 1.68 |
|  | Mean | 1.74 | 0.17 | 0.31 |
|  | Error | 1% | 2% | -4% |
| **2:Arch** | Min | 0.18 | 0.00 | 0.00 |
|  | Max | 2.14 | 0.49 | 2.01 |
|  | Mean | 0.62 | 0.19 | 0.44 |
|  | Error | 15% | 16% | -13% |
| **6:Isthmus** | Min | 0.07 | 0.04 | 0.18 |
|  | Max | 1.29 | 0.49 | 5.18 |
|  | Mean | 0.25 | 0.28 | 1.88 |
|  | Error | 56% | 43% | 44% |
| **7:Endograft** | Min | 0.07 | 0.15 | 0.31 |
|  | Max | 0.70 | 0.50 | 4.24 |
|  | Mean | 0.26 | 0.34 | 1.40 |
|  | Error | 94% | 39% | -31% |
| **8:DA** | Min | 0.08 | 0.04 | 0.07 |
|  | Max | 0.50 | 0.50 | 5.13 |
|  | Mean | 0.17 | 0.37 | 2.35 |
|  | Error | 10% | 33% | -17% |
| **9:Vao** | Min | 0.09 | 0.00 | 0.00 |
|  | Max | 13.35 | 0.49 | 3.77 |
|  | Mean | 0.56 | 0.14 | 0.65 |
|  | Error | 1% | 29% | 12% |
| **14:AbAo** | Min | 0.12 | 0.01 | 0.04 |
|  | Max | 0.43 | 0.47 | 3.05 |
|  | Mean | 0.23 | 0.21 | 0.96 |
|  | Error | 30% | 21% | 13% |

**Fig 8.**
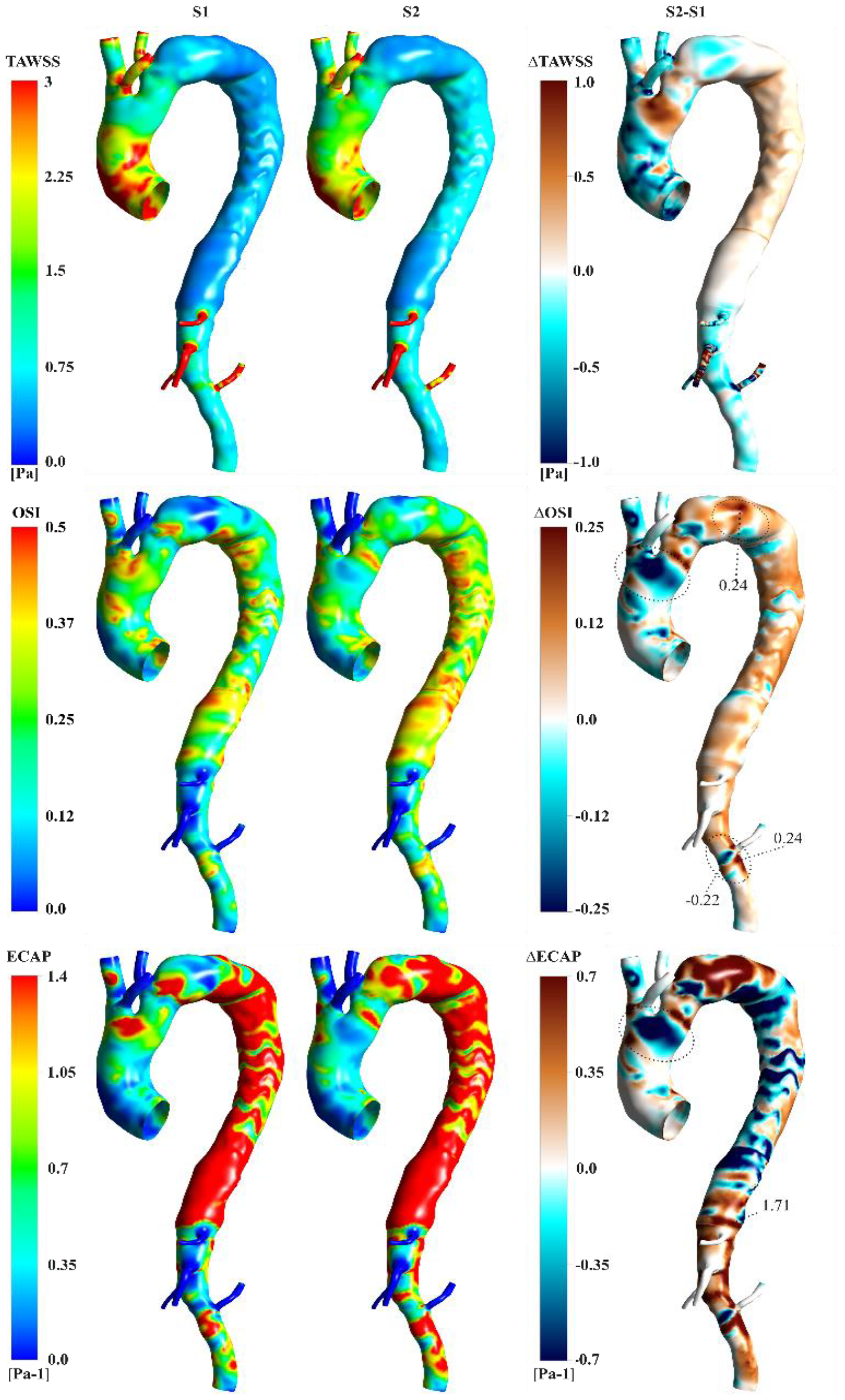
Contours of TAWSS, OSI, RRT and ECAP for S1 (left) and S2 (centre) and point-wise differences such as S1-S2 (right). Values on regions discussed in the core text are reported with dotted circles and lines.

TAWSS distributions obtained with S1 and S2 are qualitatively similar. In S2, the highest TAWSS values were found at the AA, arch and especially Vao (13.35Pa), where high velocities occur (Chen et al., 2013a; Wee et al., 2018). The OSI distributions differ broadly between S2 and S1, indicating mean relative errors >20% downstream of the arch, promoted by substantially different flow conditions predicted therein. Point-wise differences reach a min of -0.24 at the isthmus and a min and max of -0.22 and 0.24 at the Vao.

While qualitative agreements between simulations were observed in the ECAP distributions, notable differences in region exceeding the critical threshold of 1.4*Pa*^−1^ (Di Achille et al., 2014) were present in the sinotubular junction, isthmus and AbAo regions (Fig 8).

## 4. Discussion

We present a novel simulation framework driven by a single image modality (4DMRI) and informed by routinely used PWV measurements applied to a patient-specific, compliant CFD simulation of TBAD after TEVAR. Two simulation approaches were compared, differing in how the local wall stiffness informing the MBM is calculated: RPWV-based (S2) and area-based (S1). Comparing S1 and S2 simulation results highlighted that it is possible to obtain very good agreement with clinical measurements when using the RPWV-based distensibility approach whilst showing that relying solely on the area-based distensibility may not be sufficient to accurately capture patient-specific haemodynamics.

Accurate simulation of RPWVs is crucial for evaluating the haemodynamic impact of increased aortic wall stiffness after TEVAR. In this study, aortic wall stiffness, coupled with the spatial resolution limits of 4DMRI, hinders the measurement of smaller changes in aortic luminal area. Consequently, the aortic wall was considered stiff in S1, downstream of the arch, leading to inaccuracies in simulated RPWVs (Fig 4). As explained in the results section, iterative tuning of specific stiffness in S2 successfully matched in vivo RPWVs, except for the graft. A similar iterative method was employed in a Marfan syndrome CFD study using FSI, where the Young’s modulus was iteratively tuned to align the simulated PWV with 4DMRI PWV (Pons et al., 2020).

After TEVAR, a compliance mismatch is introduced between the device and the proximal aorta (Sultan et al., 2022). At the interface of the endograft, different radial displacements will occur (Cao et al., 2022), resulting in increased pressure gradient and disturbed flow dynamics. A study on flow abnormalities post-TEVAR showed an increased SFRR after the treatment (Gil-Sala et al., 2021). This is well demonstrated at the endograft-vessel interfaces where limited device dilation capacity leads to flow reversal ranges of 27-42% (Fig. 5). SFRR was substantially underpredicted in the stiff regions of S1, failing to indicate flow abnormalities.

A reduction in vorticity in the aorta has been associated with promoting aortic growth and dilation (Gallo et al., 2012; Guala et al., 2019; Stokes et al., 2023a). The IRF metric has been found to drastically reduce after TEVAR in 19 patients (Guala et al., 2020). In the present study, the IRF was overpredicted in S1 compared to S2 by up to 60%, which would indicate higher risks of rupture and would be misleading (Gil-Sala et al., 2021).

High TAWSS is triggered in regions characterised by elevated velocities and risks of local disruption (Chen et al., 2013b). Our findings revealed that the TAWSS distributions of S2 and S1 were similar. Regions of TAWSS >3Pa were found at the AA, supra-aortic and visceral branches. It is worth noting that the TAWSS distribution at the AA in S2 presents fewer regions with high values >3Pa and, thus, indicate a lower potential risk of rupture.

Collocated regions of low TAWSS and high OSI are correlated with high ECAP and aneurysmal growth (Liu et al., 2019; Zhu et al., 2021). The differences in OSI distribution observed between the simulations also led to disparities in ECAP distribution. Notably, at the isthmus, where the mean ECAP is 1.88*Pa*^−1^ and exceeds the critical threshold for S2, when this is not depicted in S1 (Fig 8.) Such disparities have significant implications for predicting thrombosis and potential vascular growth.

Additionally, the MBM assumes a linear elastic model and does not simulate longitudinal displacements, primarily in the AA (Morrison et al., 2009). However, as observed in Fig 2, the specific stiffness was reduced at the AA to match the RPWV. This allowed the longitudinal compliance to be considered within the radial compliance range and to simulate an accurate RPWV at the AA.

The limited temporal and spatial resolutions did not allow the measurement of the RPWV in any of the branches. The specific stiffness of each branch was tuned along with the proximal region to which it is attached. This assumption did not affect the simulation, as S2 matched the RPWVs well.

The work was performed with data from a single patient to build a methodology to better simulate the *in vivo* pulse wave propagation in the aorta. Future work will include validating the methodology in a patient cohort.

## 5. Conclusion

This new framework, utilising 4DMRI, significantly enhanced patient-specific compliant CFD simulations of arteries and was applied to a TBAD post-TEVAR. The comparison between simulations highlighted the accuracy of RPWV-based simulation in mimicking *in vivo* haemodynamics. The compliance mismatch introduced by TEVAR was well-captured, revealing pronounced SFRR and reduced IRF at endograft interfaces. This framework provided more reliable insights into predicting potential zones of cell deposition using WSS indices, which is crucial for evaluating aortic wall degeneration risks.

This approach represents a significant advancement, as simulation using area-based distensibility significantly differs from *in vivo* 4DMRI. RPWV-based simulation ensures more accurate haemodynamic assessments post-TEVAR, offering crucial insights for informed clinical decisions and risk predictions.

## Data Availability

All data produced in the present study are available upon reasonable request to the authors

## Acknowledgement

We thank the Department of Mechanical Engineering at University College London, the Wellcome EPSRC Centre for Interventional Surgical Sciences (WEISS) (203145Z/16/Z), the British Heart Foundation (NH/20/1/34705), the Biotechnology and Biological Sciences Research Council (BBSRC) and UK Research and Innovation (UKRI) (BB/X005062/1) for their funding support.

