## Supplemental materials for "Patient-specific compliant simulation framework informed by 4DMRI-extracted Pulse Wave Velocity: Application post-TEVAR"

### 1. Mesh independence study

In this study, relevant haemodynamics metrics were compared on three refined meshes using the Grid Convergence Index (GCI) as a validation [1]. The meshes were generated using Fluent Mesh (Ansys Inc., USA) using a size field based on curvature and proximity, a growth rate of 1.2 and maximum and minimum cell sized reported in Table 1. Ten prism layers were utilised, with the initial layer thickness set to achieve a $y+\sim1$, ensuring an accurate representation of the turbulent boundary layer. A medium mesh, denoted as M2, with maximum and minimum cell sizes of 1.5 mm and 0.5 mm, respectively, was included in a mesh independence study. M2 was compared with a coarser mesh, M1, and a finer mesh, M3. M1 and M3 were generated by approximately doubling and halving the mesh element sizing, respectively. The ratio of element counts between M1 and M2, as well as between M2 and M3, is approximately 50%. Detailed information on the element counts for each mesh is provided in Table 1:

**Table 1** Element count of M1, M2 and M3 and % change between coarse/medium and medium/fine

| | **Mesh** | | | **Change** | |
|---|---|---|---|---|---|
| | M1 | M2 | M3 | M1/M2 | M2/M3 |
| **Element Count** | 838452 | 1761218 | 3393476 | 47.6% | 51.9% |
| **Maximum Element Size [mm]** | 0.8 | 0.5 | 0.25 | | |
| **Minimum Element Size [mm]** | 2 | 1.5 | 0.8 | | |

The quality of the mesh and the analysis were assessed on seven planes and regions of interest ( see Figure 1). Mean and maximum velocities were extracted on the planes, and the mean time-averaged wall shear stress (TAWSS) was extracted on each region. The relative error between the metrics was computed between M1 and M2 and M2 and M3. Also, the GCI was computed following the study of Craven et al., [1], and calculated as follows:

$$r\sim(\frac{N_3}{N_2})^{1/3}\sim(\frac{N_3}{N_2})^{1/3} \quad (1)$$

$$p=\frac{\ln(\frac{|f_1-f_2|}{|f_2-f_3|}}{\ln(r)} \quad (2)$$

$$E_{2,1}=\frac{|f_1-f_2|}{f_2.(r^p-1)} \quad (3)$$

$$E_{3,2}=\frac{|f_2-f_3|}{f_3.(r^p-1)} \quad (4)$$

$$GCI_{2,1}=F_s|E_2| \quad (5)$$

$$GCI_{3,2}=F_s|E_3| \quad (6)$$

With $f_{1,2,3}$ the metric of interest for M1, M2 and M3, $N_{1,2,3}$ the number of elements of M1, M2 and M3, $f_{1,2,3}$ is the evaluated metric for each mesh, $F_S$ is a safety factor equal to 1.25 defined by Celik et al., [2] and used by Armour et al.,[3].

The relative error in the metrics of interest between M2 and M3 was less than 4.3%. Additionally, $GCI_{3,2}$ did not exceed 4.3%, which aligns with past research [1]. Hence, the medium mesh, M2, was chosen for the study and the analysis.

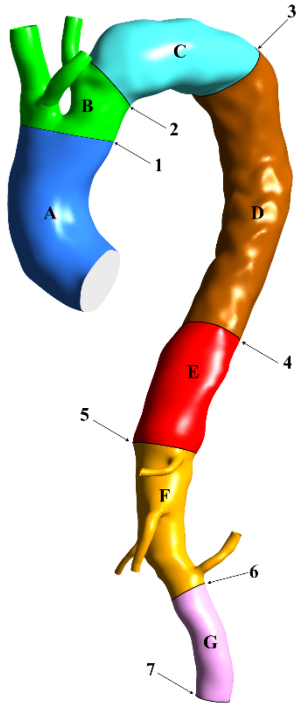


**Mean TAWSS**

| Region | A | B | C | D | E | F | G |
|---|---|---|---|---|---|---|---|
| **M1 [Pa]** | 0.661 | 0.578 | 0.292 | 0.233 | 0.220 | 2.240 | 0.590 |
| **M2 [Pa]** | 0.721 | 0.538 | 0.162 | 0.134 | 0.108 | 0.558 | 0.228 |
| **M3 [Pa]** | 0.731 | 0.558 | 0.168 | 0.139 | 0.103 | 0.542 | 0.227 |
| **%M2,M1** | 9.1% | -6.9% | -44.4% | -42.4% | -50.9% | -75.1% | -61.4% |
| **%M3,M2** | 1.4% | 3.7% | 3.6% | 3.6% | -4.6% | -2.9% | -0.5% |
| **GCI 2,1** | 1.8% | 9.0% | 5.1% | 4.8% | 6.0% | 3.7% | 0.6% |
| **GCI 3,2** | 0.1% | 4.3% | 0.5% | 0.2% | 0.3% | 0.0% | 0.0% |

**Mean velocity**

| Plane | 1 | 2 | 3 | 4 | 5 | 6 | 7 |
|---|---|---|---|---|---|---|---|
| **M1 [m/s]** | 0.078 | 0.067 | 0.021 | 0.023 | 0.017 | 0.059 | 0.058 |
| **M2 [m/s]** | 0.082 | 0.059 | 0.032 | 0.036 | 0.026 | 0.048 | 0.053 |
| **M3 [m/s]** | 0.0840 | 0.0610 | 0.0310 | 0.0350 | 0.0270 | 0.0500 | 0.0510 |
| **%M2,M1** | 5.1% | -11.9% | 50.5% | 56.5% | 54.7% | -19.3% | -9.0% |
| **%M3,M2** | 2.4% | 3.4% | -1.9% | -2.8% | 2.7% | 5.0% | -3.4% |
| **GCI 2,1** | 6.1% | 5.6% | 2.5% | 3.8% | 3.6% | 8.0% | 6.5% |
| **GCI 3,2** | 3.0% | 1.4% | 0.1% | 0.3% | 0.3% | 1.6% | 2.3% |

**Max Velocity**

| Plane | 1 | 2 | 3 | 4 | 5 | 6 | 7 |
|---|---|---|---|---|---|---|---|
| **M1 [m/s]** | 0.256 | 0.25 | 0.171 | 0.168 | 0.175 | 0.271 | 0.257 |
| **M2 [m/s]** | 0.281 | 0.343 | 0.267 | 0.218 | 0.157 | 0.244 | 0.207 |
| **M3 [m/s]** | 0.285 | 0.346 | 0.259 | 0.224 | 0.162 | 0.252 | 0.209 |
| **%M2,M1** | 9.8% | 37.2% | 56.1% | 29.8% | -10.3% | -10.0% | -19.5% |
| **%M3,M2** | 1.4% | 0.9% | -3.0% | 2.8% | 3.2% | 3.3% | 1.0% |
| **GCI 2,1** | 2.1% | 1.1% | 4.1% | 3.9% | 5.5% | 5.8% | 1.3% |
| **GCI 3,2** | 0.3% | 0.0% | 0.4% | 0.5% | 1.5% | 1.7% | 0.0% |

**Figure 1: On the left side are planes and regions of interest, denoted by numbers and capital letters**. On the right side, tables display the relative error and GCI comparison for M1, M2, and M3 regarding the mean TAWSS and mean and maximum velocity.

### 2. Three Element Windkessel Parameters -outlet boundary conditions

Below is a table grouping the three-element Windkessel parameters used at the outlets of the aorta to apply a pressure condition mimicking the effects of the peripheral vasculature system. The abbreviations stand for right common carotid (RCC), right subclavian (RSA), left subclavian (LSA), coeliac trunk (CT), superior mesenteric (SMA), right renal (RR), left renal (LR), and abdominal aorta (AbAo). Considering the compliance of the aorta downstream of the arch in S2 after the specific stiffness leads to an increase in aortic compliance. As the total compliance does not vary, the peripheral compliance and Windkessel compliance were proportionally decreased in S2 [4].

**Table 1** Three-element Windkessel parameters for S1 and S2. R1 and R2 are in [mmHg.mL.s] and C is in [mL/mmHg].

| | | RCC | RSA | LCC | CT | SMA | LR | RR | AbAo |
|---|---|---|---|---|---|---|---|---|---|
| **S1** | **R1** | 0.33 | 0.62 | 0.57 | 1.47 | 0.55 | 12.31 | 15.33 | 0.31 |
| | **R2** | 4.03 | 10.47 | 9.57 | 24.82 | 9.19 | 31.65 | 39.42 | 5.29 |
| | **C** | 0.143 | 0.056 | 0.061 | 0.024 | 0.062 | 0.014 | 0.011 | 0.111 |
| **S2** | **R1** | 0.35 | 0.65 | 0.59 | 1.54 | 0.57 | 12.89 | 16.06 | 0.33 |
| | **R2** | 4.22 | 10.97 | 10.02 | 26.00 | 9.63 | 33.15 | 41.29 | 5.54 |
| | **C** | 0.037 | 0.014 | 0.016 | 0.006 | 0.016 | 0.004 | 0.003 | 0.029 |
